# Performance of three SARS-CoV-2 immunoassays, three rapid lateral flow tests and a novel bead-based affinity surrogate test for the detection of SARS-CoV-2 antibodies in human serum

**DOI:** 10.1101/2021.02.07.21251062

**Authors:** Manuel Krone, Julia Gütling, Johannes Wagener, Thiên-Trí Lâm, Christoph Schoen, Ulrich Vogel, August Stich, Florian Wedekink, Jörg Wischhusen, Thomas Kerkau, Niklas Beyersdorf, Silvana Klingler, Simone Backes, Lars Dölken, Georg Gasteiger, Oliver Kurzai, Alexandra Schubert-Unkmeir

## Abstract

For the control of immunity in COVID-19 survivors and vaccinated subjects there is an urgent need for reliable and rapid serological assays.

Based on samples from 63 COVID-19 survivors up to seven months after symptom onset, and on 50 serum samples taken before the beginning of the pandemic, we compared the performance of three commercial immunoassays for the detection of SARS-CoV-2 IgA and IgG antibodies (Euroimmun SARS-COV-2 IgA/IgG, Mikrogen *recomWell* SARS-CoV-2 IgA/IgG, and SERION ELISA *agile* SARS-CoV-2 IgA/IgG) and three rapid lateral flow (immunochromatographic) tests (Abbott Panbio COVID-19 IgG/IgM, NADAL COVID-19 IgG/IgM, and Cleartest Corona 2019-nCOV IgG/IgM) with a plaque-reduction neutralization test (PRNT50) representing the gold standard. In addition, we report and validate a novel, non-commercial flow cytometry bead-based surrogate test.

57 out of 63 PCR-confirmed COVID-19 patients (90 %) showed neutralizing antibodies. The sensitivity of the seven assays ranged from 7.0 % to 98.3 %, the specificity from 86.0 % to 100.00 %. Only one commercial immunoassay showed a sensitivity and specificity of greater than 98 %. These data indicate abundant interassay variability.

## Introduction

The novel severe acute respiratory syndrom coronavirus 2 (SARS-CoV-2) causing coronavirus disease (COVID-19) was first reported in China at the end of 2019 and since then has caused a worldwide pandemic. As of February 7, 2021, more than 105 million cases and 2.2 million deaths have been reported attributed to COVID-19 worldwide (1).

SARS-CoV-2 is mainly transmitted from person to person through respiratory droplets and aerosols, although indirect transmission via contaminated objects or surfaces may also be possible (2, 3). The virus enters host cells via the angiotensin-converting enzyme 2 (ACE2) (4, 5), which is highly expressed on lung alveolar epithelial cells and small intestinal epithelial cells (6). Typical clinical symptoms of COVID-19 are olfactory or gustatory dysfunction, fever, fatigue, dry cough, shortness of breath, headache, and joint pain. Severe COVID-19 cases can result in acute respiratory distress syndrome, septic shock, metabolic acidosis, coagulation dysfunction, and multiple organ dysfunction syndromes (7). In order to control the spread of the virus, many countries have implemented stringent non-pharmaceutical interventions as well as extensive laboratory diagnostic tests of symptomatic but also asymptomatic people. A rapid and early laboratory diagnosis is critical to diagnose infection and control transmission. As the gold standard test for SARS-CoV-2 identification, direct pathogen detection by nucleic acid amplification tests (NAAT), such as real-time RT-PCR, is the method of choice both for patient detection and outbreak management (8, 9). In addition, rapid antigen testing is used in point-of-care diagnostics. Based on the currently available studies, the sensitivity of ELISA assays for antibody detection increases during the second week after the onset of symptoms (10) when infectiosity is most likely lost and is therefore not eligible for diagnosis of active COVID-19 disease.

Nevertheless, antibody testing is needed to determine if persons have been exposed to SARS-CoV-2, in patients with potential post-COVID-19-symptoms as well as epidemiological investigations. In patients with doubtful PCR results, antibody testing helps to distinguish between active SARS-CoV-2 infections, persistant PCR positivity after an undiagnosed SARS-CoV-2 infection or false positive PCR results (11). It also allows the identification of suitable convalescent plasma donors and may become important for the determination of post-vaccination immunity. For these purposes, especially a high specificity is important to avoid “false immune” cases.

Virus neutralization assays remain the gold standard for determining antibody efficacy but are complex, difficult to standardize, and time-consuming. In contrast, enzyme-linked immunosorbent assays (ELISA) are feasible in a routine laboratory setting and cheap. To support decision making on the employment of antibody testing for either diagnostic or population screening, we present a detailed comparison of serological COVID-19 assays to virus neutralization assays.

Several publications have assessed sensitivities of various ELISAs for laboratory use and lateral flow assays for point-of-care use as proportion of previously PCR positive patients at least 14 days after the PCR and found sensitivities between 15 % to 100 % (12-15), but only few have evaluated these assays against tests that directly measure virus neutralization (16-19).

The aim of this study was to evaluate the performance and utility for routine diagnostic testing of three commercially available immunoassays [Euroimmun SARS-COV-2 IgA and IgG (CE marked), Mikrogen *recomWell* SARS-CoV-2 IgA and IgG (CE marked) and SERION ELISA *agile* SARS-CoV-2 IgA and IgG (CE marked)], three rapid lateral flow (immunochromatographic) tests [Abbot Panbio COVID-19 IgG/IgM rapid test device (CE marked), NADAL COVID-19 IgG (CE marked) and Cleartest Corona 2019-nCOV IgG (CE marked)], and a non-commercial bead-based surrogate test designed to estimate antibody affinity and thereby better predict SARS-CoV-2 neutralizing activity in comparison to a pseudotype-based virus (micro)neutralization assay (20) that has recently been shown to accurately measure protective immunity against SARS CoV-2 (21).

## Material and Methods

### Study Protocol

Patients participating in the study were asked to answer a questionnaire about their previous COVID-19 disease and serum specimens were collected from the participants. Inclusion criteria were a PCR-confirmed COVID-19 disease with a first positive SARS-CoV-2-PCR at least 14 days before serum collection, age of at least 18 years and a written informed consent. In patients with more than one available serum specimen, the first specimen fulfilling inclusion criteria was included into the main analysis.

### Questionnaire

All participants filled out a questionnaire including questions about age, gender, date of symptom onset, date of first positive PCR test, symptoms (fever, cough, sore throat, dyspnea, rhinorrhea, headache, joint pain, diarrhea, olfactory or gustatory dysfunction, other), comorbidities (chronical pulmonary disease, ischemic heart disease, autoimmunological disease) as well as immunodeficiencies, intake of corticosteroids (systemically or inhaled) and other immunosuppressants.

### Serum samples comprising the negative panel

All serological assays were performed on 50 serum samples from 50 different individuals. All of these samples were collected in Spring 2018, before the circulation of SARS-CoV-2 in Europe. Use of this samples was approved by the local ethics committee.

### Serum samples comprising the positive COVID-19 patient panel

Serum samples from the participants were collected. The date of symptom onset, clinical classification and basic demographic information (male/female, age) were recorded.

### Pseudotyping of Vesicular Stomatitis Virus with SARS-CoV-2 Spike protein

Pseudotyping of Vesicular Stomatitis Virus (VSV) with the SARS-CoV2 Spike protein was performed as previously described (5). Briefly, VSVΔG-eGFP/luciferase was pseudotyped on BHK cells inducibly expressing the VSV G protein (kindly provided by Gert Zimmer) to generate VSVΔG-G. Next, 293T cells were transfected with a plasmid expressing SARS-CoV2 Spike (kindly provided by Markus Hoffmann and Stefan Pöhlmann, (5)) using TransIT-X2 (Mirus) according to the manufacturer’s instructions. 24 hours post transfection, cells were infected with VSVΔG-G. In order to neutralize residual input virus, inoculated cells were washed twice with PBS two hours post infection and new medium containing 1:1000 anti-VSV-G antibody (8G5F11, Kerafast) was added. Supernatant containing replication-deficient VSVΔG pseudotyped with SARS-CoV2 Spike protein (VSVΔG-S) was harvested 18 hours post infection, clarified by centrifugation and stored at -80°C. Viral titers were determined on Vero E6 cells by the quantification of green fluorescence.

### Pseudovirus neutralisation assay

For pseudovirus neutralisation assays, Vero cells (ATCC CCL-81) were seeded in 96-well plates (20.000 cells/plate) in culture medium and were incubated at 37°C for 4 hours. Sera were serially diluted 1:3 in infection medium starting with a 1:10 dilution. VSVΔG-S pseudoparticles were diluted in infection medium for a GFP+ cell count in the assay of ∼200/well. Serum dilutions were mixed 1:1 with pseudoparticles and incubated for 45 minutes at 37°C prior to addition to the preplated Vero cells and incubation at 37 °C for 16 hours. The number of GFP+ viral foci was counted using a fluorescent microscope. The 50% pseudovirus neutralization titre (pVNT50) was reported as the interpolated reciprocal of the dilution yielding a 50% reduction in fluorescent viral foci. Neutralization against GFP-expressing recombinant VSV was assayed in parallel to exclude unspecific inhibition of infection. In case of an inhibition greater than 30 % in the inhibition control, PRNT50 titers were read in relation to the PFU in the inhibition control. Neutralization testing was repeated two times in sera with a SARS-CoV-2 independent neutralization greater than 50 % as well as in sera negative in the neutralization testing but with at least one borderline or positive ELISA test. The median titer was then selected for further analysis. PRNT50 ≥ 20 were considered positive.

### Immunoassays for anti-SARS-CoV-2 IgA and IgG

ELISA kits from three manufacturers were included in the study: Euroimmun SARS-COV-2 IgA and IgG (CE marked, Euroimmun Medizinische Labordiagnostika, Lübeck, Germany), Mikrogen *recomWell* SARS-CoV-2 IgA and IgG (CE marked, Mikrogen, Neuried, Germany) and SERION ELISA *agile* SARS-CoV-2 IgA and IgG (CE marked, Institut Virion/Serion, Würzburg, Germany). The Euroimmun assay is based on S1 antigen, Mikrogen *recomWell* is based on the nucleocapsid (N) protein as antigen and SERION ELISA *agile* is coated with both S1 and S2 and the nucleocapsid. The assays were performed following the manufacturers’ instruction. The following thresholds were used: In the Mikrogen assays, samples with a concentration of < 20 U/ml were assessed as negative, samples with a concentration of ≥ 20 U/ml but < 24 U/ml were counted as borderline, samples with a concentration of ≥ 24 U/ml were counted positive. For Euroimmun’s IgA and IgG assays, extinction ratios were calculated out of specimen extinction and calibrator extinction. Ratio values < 0.8 were considered as negative; those with a ratio ≥ 1.1 were considered as positive and those with a ratio ≥ 0.8 to < 1.1 were recorded as borderline according to the manufacturer’s recommendations. The thresholds in the SERION assay were < 10 U/ml for negative results, ≥ 10 U/ml but < 14 U/ml for IgA and ≥ 10 U/ml but < 15 U/ml for IgG antibodies as borderline and > 14 U/ml for IgA and > 15 U/ml for IgG as positive.

### Rapid lateral flow tests

The serum samples were also used to evaluate three rapid lateral flow antibody tests: NADAL COVID-19 IgG/IgM Test (whole blood, serum or plasma) from new art laboratories (nal)-von minden (Moers, Germany) targeting a not further specified SARS-CoV-2-Antigen, Panbio COVID-19 IgG/IgM rapid test device from Abbott (Chicago IL, USA) utilizing the nucleocapsid (N) protein (14) and the Cleartest Corona 2019-nCOV IgM/IgG (Servoprax, Wesel, Germany) targeting a “2019-nCOV-Antigen”. All three tests are based on immunochromatography for the qualitative detection of IgM and IgG antibodies specific to SARS CoV-2 in human serum, plasma, fingerstick and venipuncture whole blood. The tests were performed following the manufacturers’ instructions. Readout (positive/negative) was interpreted by two independent operators. As volume of the negative sera was limited, not all negative sera could be tested with all lateral flow tests.

### Bead-based surrogate test for the detection of neutralizing anti-SARS-CoV-2 antibodies

Dynabeads^®^ M-270 Epoxy (ThermoFisher, Waltham, USA) were coated according to the manufacturer’s instructions with different concentrations of either commercially available full-length SARS-CoV2 S1-Spike-Protein (His-tagged, Sino Biological, Beijing, China) expressed and purified from HEK cells or with SARS-CoV-2 RBD protein expressed and purified from Expi293F™ cells (22-26). Importantly, the receptor binding domain of the viral spike protein has been found to be an immunodominant and highly specific target of antibodies in SARS-CoV-2 patients (27). The coated beads were resuspended in PBS / 0.1 % BSA and stored at 4°C. For detection of anti-SARS-CoV-2 mAb, beads were first blocked with PBS / 10% BSA followed by incubation with 1:100 dilution of serum in PBS / 0.1 % BSA / 0.02% NaN3. After extensive washing, bead-bound antibodies were detected with Goat-anti-human IgG Alexa 647 (Dianova, Hamburg, Germany). A mouse/human chimeric monoclonal anti-SARS-CoV2-Spike-S1 antibody (Sino Biological, Beijing, China) was used as a positive control for all measurements. Samples were measured on a FACS Calibur flow cytometer (Becton Dickinson, Franklin Lakes NJ, USA) and FlowJo software version 10 (TreeStar, Ashland OR, USA) was used for data analysis. The flow cytometric data were converted into Scores taking into account binding and affinity of the bound antibodies.

RBD-Score: a) binding: ratio MFI(sample)/MFI(negative control) ≤ 1 score 0, > 1 ≤ 1,19 score 1, ≥1,2 ≤ 1,99 = score 2, > 1,99 score 3; b) affinity: MFI(sample – beads with low Ag coating)/MFI(sample – beads with high Ag coating): ≤ 39,99 % score 1, > 39,99 % sore 1.5; Total score(RBD) = score(binding) x score(affinity).

S1-Score: a) binding: ratio MFI(sample)/MFI(negative control) ≤ 1,19 score 0, > 1,19 ≤ 1,99 score 1, > 1,99 score 2; b) affinity: MFI(sample – beads with low Ag coating)/ MFI(sample – beads with high Ag coating): ≤ 74,99 % score 1, > 74,99 % score 2; Total score(S1) = score(binding) x score(affinity); Total score = score(RBD) x score(S1).

### Statistical analysis

Data analysis was performed using Microsoft Excel 2016, IBM SPSS Statistics 26, and GraphPad Prism 8. Differences in symptoms between patients with and without neutralizing antibodies as well as differences in test sensitivity were analyzed using two-tailed Fisher’s exact test. Differences in the time interval between PCR result and serum collection were compared using Student’s t-test. Spearman’s rank correlation coefficient rho was calculated to determine the correlation of different tests. Two-tailed significance was calculated using exact permutation based probabilities. For the calculation of specificity and sensitivity, borderline results of the serological tests were counted as positive. Specificity was calculated as the proportion of negatively detected samples in the negative sera from 2018, sensitivity was calculated as proportion of samples with neutralizing activity in the pseudovirus neutralization assay. Confidence intervals (CI) were calculated using the Wilson score method without continuity correction (28) using a Confidence interval calculator sheet (29). A significance level α of 0.05 was used.

## Ethics declaration

The study protocol was approved by the local ethics committee in accordance to the declaration of Helsinki (File No 84/20).

## Results

### Patient selection

Out of 68 participating patients, 63 were included in the study. Five patients did not meet the inclusion criteria: Three patients reported that their diagnosis was not based on a PCR result but on SARS-CoV-2 antibody rapid testing. Neutralizing antibodies were not found in these three patients, but various commercial assays were positive for SARS-CoV-2 antibodies. The sera of two patients were taken 3 and 13 days, respectively, and thus too early after their first positive PCR result.

## Demographic data

45 of the 63 patients, mostly health care workers, assigned themselves to a female gender (71 %), 18 to a male gender (29 %), no participant felt belonging to diverse gender. The gender and age distribution of the participants is shown in Fig.1.

**Fig. 1:**
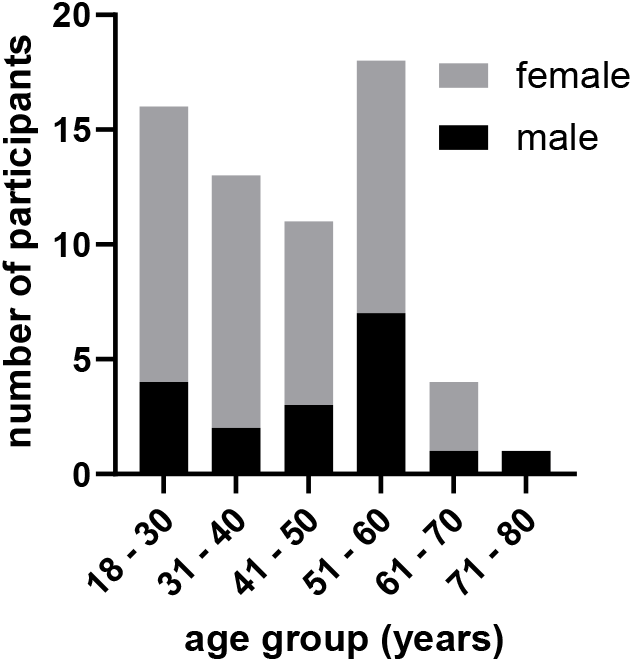
Participants’ demographics (n=63)

The median time span between the first positive PCR result and serum collection was 91 days (Range: 14 - 226 days), the median time span between the onset of symptoms and serum collection was 94 days (Range: 17 - 228 days).

### Clinical characteristics of the participants of the study

62 of 63 participants answered questions of their course of disease, comorbidities, and immunosuppression. 60 of 62 participants reported symptoms, two did not report any symptoms. The most common symptoms were olfactory or gustatory dysfunction (69 %, 43/62), headache (55 %, 34/62), cough (52%, 32/62), joint pain (44 %, 27/62), rhinorrhea (42 %, 26/62), sore throat (39 %, 24/62), dyspnea (29 %.18/62), and diarrhea (21 %, 13/62). Other reported symptoms were fatigue (16 %, 10/62), nausea (5 %, 3/62), angina pectoris (5 %, 3/62), dizziness (3 %, 2/62). Mucosal dryness, erythema, back pain, shivering, inappetence, vision disorder, and fine motor difficulties, reported by one participant each. Fever was reported by 31 of 62 patients (50 %). In 4 % (2/57) of cases, no symptoms were reported. Eight patients (13 %) reported chronical pulmonary disease, five (8 %) autoimmunological disease and one (2 %) ischemic heart disease as comorbidities. One participant had an immunodeficiency (2 %), nine (15 %) had inhaled corticosteroids; three participants had taken systemic corticosteroids during the last year. None of the participants had taken other immunosuppressant or immunomodulatory substances during the last year.

### Pseudovirus neutralisation assay

Neutralizing SARS-CoV-2 antibodies could be detected in 57 of the 63 patients. Neutralization assay results could be reproduced in all cases except one where the initial test showed no neutralization at all but both repeated measures showed neutralization (50% plaque-reduction neutralization test - PRNT50: 540). The distribution of the PRNT50 titres are shown in Fig. 2.

**Fig. 2:**
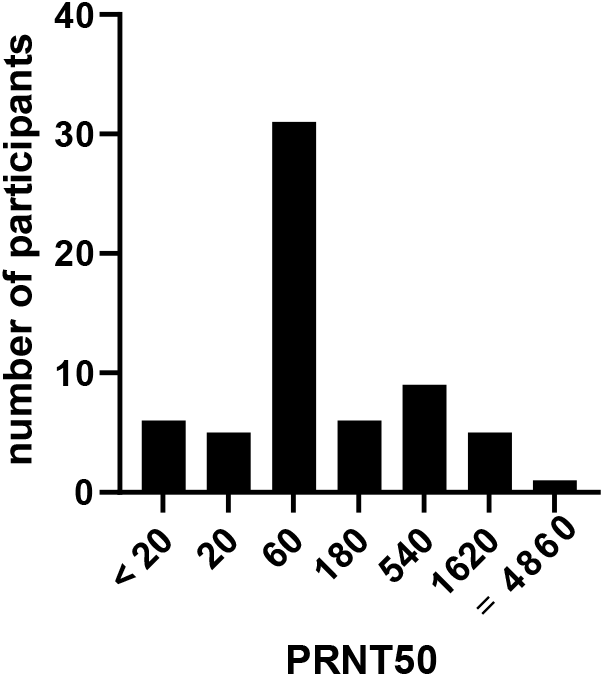
Distribution of the PRNT50 titers of the included patients (n = 63). PRNT50: 50 % plaque reduction neutralization test. Method described in the methods section. PRNT50 < 20 were considered negative.

Both asymptomatic participants were tested negative (p < 0.01). None of the patients without neutralizing antibodies reported fever, compared to 55 % (31/56) of the patients with neutralising antibodies (p = 0.02). No other significant differences in symptoms could be determined between patients with and without neutralizing antibodies. The median time between first PCR and serum collection (median: 82 days, minimum: 26 days, maximum: 194 days) did not differ significantly from the patients with neutralizing antibodies (p = 0.79). Neutralizing antibodies could not be detected in any of the 50 sera from 2018.

### Comparison of commercial immunoassays with PRNT50

Results of Euroimmun SARS-COV-2 IgA/IgG, Mikrogen *recomWell* SARS-CoV-2 IgA/IgG and SERION ELISA *agile* SARS-CoV-2 IgA/IgG were plotted against the titers observed in the neutralization assays (Fig. 3).

**Fig. 3:**
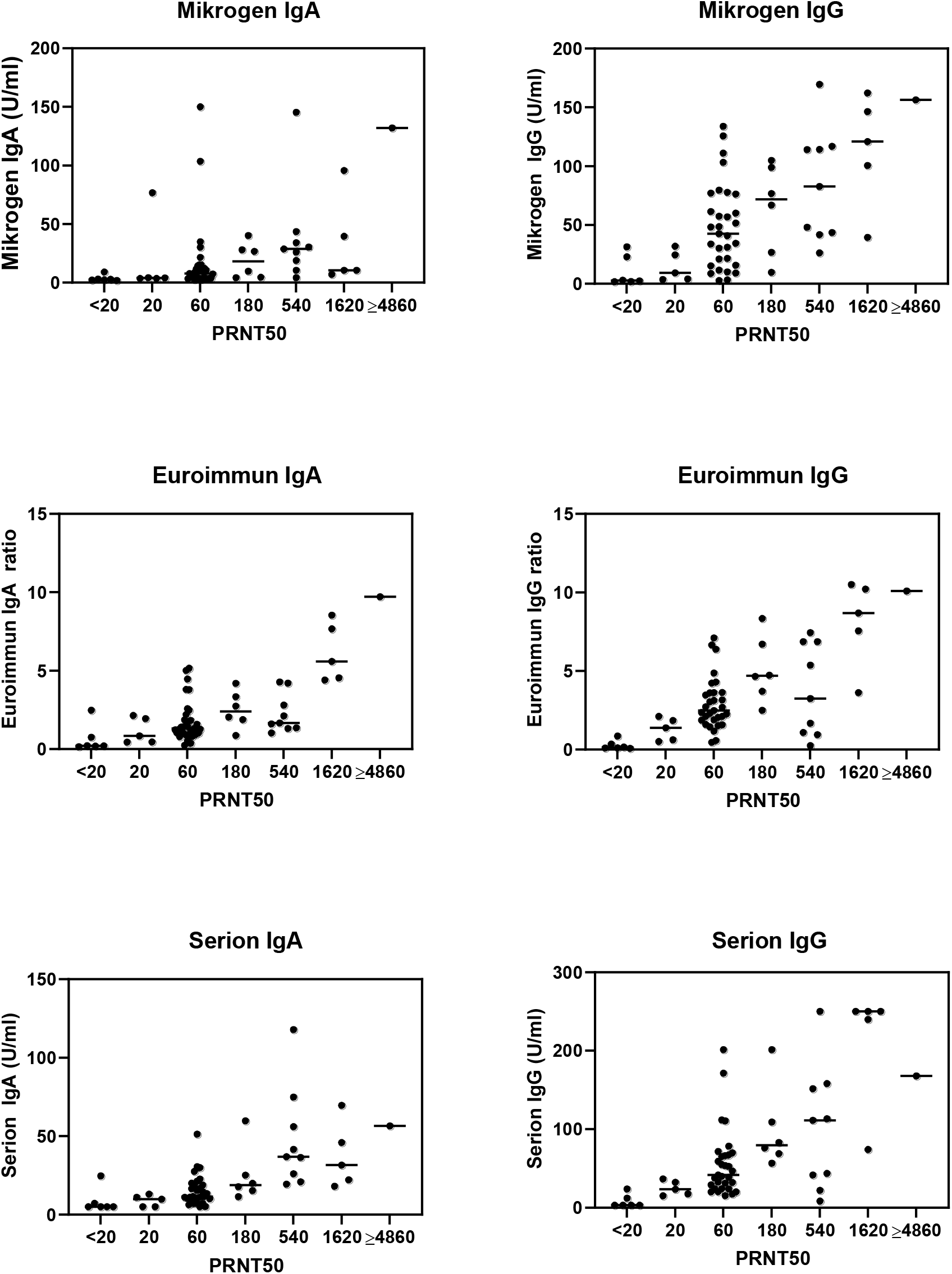
Results of the ELISA assays plotted against the titers from the neutralization assay (n=63). Values exceeding the maximum respectively the minimum in a test were plotted at this maximum or minimum value (Mikrogen IgA: >150.12 U/ml; Mikrogen IgG: >133.93 U/ml; Euroimmun IgA: >8.54; SERION IgA: <5.0 U/ml; SERION IgG: <3.0 U/ml; >250.0 U/ml) Mikrogen: Mikrogen recomWell SARS-CoV-2 IgA / IgG (Mikrogen, Neuried, Germany), based on the nucleocapsid Euroimmun: Euroimmun SARS-COV-2 IgA / IgG (Euroimmun, Lübeck, Germany), based on S1 antigen SERION: SERION ELISA agile SARS-CoV-2 IgA / IgG (Institut Virion/Serion, Würzburg, Germany), based on the nucleocapsid as well as S1 and S2 antigen PRNT50: 50 % plaque reduction neutralization test. Method described in the methods section. PRNT50 < 20 were considered negative.

The sensitivities of the IgA assays to detect neutralizing antibodies ranged from 31.6 % (Mikrogen) to 91.2 % (Euroimmun), the specificities from 86.0 % (Euroimmun) to 100.0 % (SERION). The sensitivities of the IgG assays ranged from 79.0 % (Mikrogen) to 100.0 % (SERION), the specificities from 94.0 % (Mikrogen) to 100.0 % (SERION). Detailed data and confidence intervals are shown in Table 1.

**Table 1:** Sensitivity and Specificity data of the six ELISA assays included in the study.

| Assay | Sensitivity (95% CI) | Specificity (95% CI) |
| --- | --- | --- |
| **Mikrogen IgA** | 31.6 % (21.0 – 44.5 %) | 94.0 % (83.8 – 97.9 %) |
| **Mikrogen IgG** | 79.0 % (66.7 – 87.5 %) | 94.0 % (83.8 – 97.9 %) |
| **Euroimmun IgA** | 91.2 % (81.1 – 96.2 %) | 86.0 % (73.8 – 93.1 %) |
| **Euroimmun IgG** | 91.2 % (81.1 – 96.2 %) | 96.0 % (86.5 – 98.9 %) |
| **SERION IgA** | 82.5 % (70.6 – 90.2 %) | 100.0 % (92.9 – 100.0 %) |
| **SERION IgG** | 98.3 % (90.7 – 99.7 %) | 100.0 % (92.9 – 100.0 %) |

*Sensitivity data were collected on 57 samples positive in the neutralization assay. Specificity data were collected on 50 negative sera.*
*Confidence intervals were calculated using the Wilson score method without continuity correction (28) using a Confidence interval calculator sheet (29).*
*Mikrogen: Mikrogen recomWell SARS-CoV-2 IgA / IgG (Mikrogen, Neuried, Germany)*
*Euroimmun: Euroimmun SARS-COV-2 IgA / IgG (Euroimmun, Lübeck, Germany)*
*SERION: SERION ELISA agile SARS-CoV-2 IgA / IgG (Institut Virion/Serion, Würzburg, Germany)*
*95% CI: 95% Confidence Interval*

While the sensitivities of Mikrogen IgA (p < 0.001), Mikrogen IgG (p = 0.002) and SERION IgA (p = 0.008) were significantly lower compared to the best performing assay SERION IgG, the differences between Euroimmun IgA / IgG and SERION IgG were not significant (p = 0.21). All ELISA values correlated significantly with the PRNT50 values (p < 0.001). rho values ranged from 0.562 (Mikrogen IgA) to 0.685 (SERION IgG) and 0.715 (SERION IgA).

A combination of IgG and IgA (IgA or IgG) positivity increases the sensitivity of SERION to 100.0 % (95% CI: 92.9 – 100.0 %) without specificity losses. The combined sensitivity for Euroimmun IgG and IgA increases to 96.5 % (95% CI: 88.1 – 99.0 %) with a loss of specificity to 86.0 % (95% CI: 73.8 – 93.1 %). In Mikrogen no increase in sensitivity could be reached combining IgG and IgA, but specificity decreases to 88.0 % (95% CI: 76.2 – 84.4 %).

### Rapid antibody testing (Lateral flow assays)

The sensitivities of the IgM assays to detect neutralizing antibodies ranged from 7.0 % (Abbott) to 66.7 % (NADAL), the specificities from 91.1 % (Cleartest) to 100.0 % (Abbott and NADAL). The sensitivities of the IgG assays ranged from 84.2 % (Abbott) to 89.5 % (Cleartest), the specificities from 95.6 % (Cleartest) to 100.0 % (Abbott and NADAL). Detailed data and confidence intervals are shown in Table 2. Regarding sensitivity, all rapid antibody tests were significantly inferior to the SERION IgG ELISA regarding sensitivity (p = 0.03 and lower) except for the Cleartest IgG where statistical significance was not reached (p = 0.11).

**Table 2:** Sensitivity and Specificity data of the six lateral flow immunochromatographic assays included in the study.

| Assay | Sensitivity (95%CI) | Specificity (95%CI) |
| --- | --- | --- |
| Abbott IgM | 7.0 % (2.8 – 16.7 %) | 100.0 % (92.4 – 100.0 %) |
| Abbott IgG | 84.2 % (72.6 – 91.5 %) | 100.0 % (92.4 – 100.0 %) |
| NADAL IgM | 66.7 % (53.7 – 77.5 %) | 100.0 % (92.1 – 100.0 %) |
| NADAL IgG | 86.0 % (74.7 – 92.7 %) | 100.0 % (92.1 – 100.0 %) |
| Cleartest IgM | 21.1 % (12.5 – 33.3 %) | 91.1 % (79.3 – 96.5 %) |
| Cleartest IgG | 89.5 % (78.9 – 95.1 %) | 95.6 % (85.2 – 98.8 %) |

Sensitivity data were collected on 57 samples positive in the neutralization assay. Specificity data were collected on 47
*negative sera (Abbott) or 45 negative sera (NADAL and Cleartest).*
Confidence intervals were calculated using the Wilson score method without continuity correction (28) using a Confidence interval calculator sheet (29).
Abbot: Abbott Panbio COVID-19 IgM / IgG (Abbott, Chicago IL, USA)
NADAL: NADAL COVID-19 IgM / IgG (nal von minden, Moers, Germany)
*Cleartest: Cleartest Corona 2019-nCoV IgM / IgG (Servoprax, Wesel, Germany)*
95% CI: 95% Confidence Interval

**Table 3:** Antibody testing results in one individual in chronological sequence. The first positive PCR sample was taken the day after symptom onset.

| Days after onset of symptoms | PRNT50 | Mikrogen IgA (U/ml) | Mikrogen IgG (U/ml) | Euroimmun IgA (ratio) | Euroimmun IgG (ratio) | SERION IgA (U/ml) | SERION IgG (U/ml) | BBST (Total Bead Score) |
| --- | --- | --- | --- | --- | --- | --- | --- | --- |
| 2 days | <20 | NEG (3.25) | NEG (3.02) | NEG (0.48) | NEG (0.20) | NEG (<5.00) | NEG (<3.00) | NEG (0) |
| 5 days | <20 | NEG (3.12) | NEG (3.40) | NEG (0.47) | NEG (0.20) | NEG (<5.00) | NEG (<3.00) | NEG (0) |
| 8 days | <20 | NEG (4.75) | NEG (3.63) | NEG (0.55) | NEG (0.22) | POS (29.21) | NEG (<3.00) | NEG (0) |
| 11 days | 60 | POS (48.64) | POS (25.84) | POS (1.34) | NEG (0.37) | POS (131.54) | POS (17.84) | NEG (0) |
| 14 days | ≥4860 | POS (56.86) | POS (39.12) | POS (1.48) | NEG (0.57) | POS (148.26) | POS (22.50) | POS (3) |
| 17 days | 540 | POS (43.59) | POS (41.68) | POS (1.32) | BOR (0.94) | POS (117.97) | POS (22.20) | NEG (0) |
| 24 days | 540 | POS (25.93) | POS (44.72) | BOR (0.78) | POS (1.16) | POS (66.21) | POS (18.37) | POS (6) |
| 121 days | 60 | NEG (5.13) | NEG (18.95) | POS (1.16) | POS (2.87) | POS (22.78) | POS (17.55) | POS (9) |
| 174 days | 60 | NEG (4.46) | NEG (10.81) | BOR (0.92) | POS (2.79) | POS (20.44) | POS (17.42) | NEG (0) |

*PRNT50: 50 % plaque reduction neutralization test. Method described in the methods section.* *Euroimmun: Euroimmun SARS-COV-2 IgA / IgG (Euroimmun, Lübeck, Germany)* *Mikrogen: Mikrogen recomWell SARS-CoV-2 IgA / IgG (Mikrogen, Neuried, Germany)* *SERION: SERION ELISA agile SARS-CoV-2 IgA / IgG (Institut Virion/Serion, Würzburg, Germany)* *BBST: Bead-based surrogate test. Method described in the methods section.*

### Bead-based surrogate test

The sensitivity of the bead-based surrogate test for neutralizing antibodies was 71.9 % (95% CI: 59.2 – 81.9 %), the specificity 100 % (95 % CI: 92.9 – 100.0 %). The total bead score is correlated with the inhibition corrected PRNT50 (rho = 0.414; p = 0.001, Fig. 4).

**Fig. 4:**
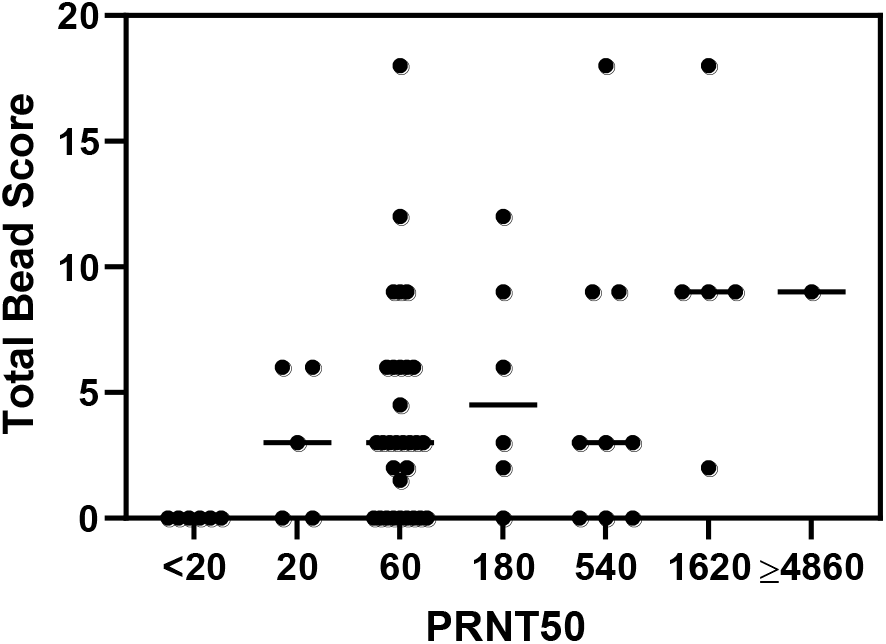
Results of the bead-based surrogate test plotted against the titers from the neutralization assay (n=63). PRNT50: 50 % plaque reduction neutralization test. Method described in the methods section. PRNT50 < 20 were considered negative.

### Survivors of a PCR confirmed COVID-19 disease without neutralizing antibodies

In three of the six patients without detectable neutralizing antibodies against the full-length Spike protein or the Spike protein receptor-binding domain, no IgM, IgA or IgG antibodies could be detected in any of the six commercial assays. In one patient, Mikrogen and SERION showed borderline results in the IgG assay and the Abbott IgG showed a positive result, in one patient only Nadal IgM/IgG was positive. Interestingly, one patient had positive IgG antibodies in the Mikrogen, SERION, and Abbott assay as well as positive IgM and borderline IgG in the Euroimmun assay. The bead-based surrogate test was negative in all of these six patients.

### Antibody dynamic

While the IgA antibodies determined by the Mikrogen ELISA assay were significantly negatively correlated with the time between the first positive PCR and the serum sampling (rho = -0.313; p=0.01), this was not the case for antibodies measured by other ELISAs, neutralization testing and the flow cytometry bead-based test.

We followed the seroconversion against SARS-CoV-2 in one representative patient with moderate symptoms using the neutralization assay, all six ELISA systems, and the bead-based surrogate test (Tab.3): Positive IgA levels were detected from day 8 after onset of symptoms with the SERION IgA ELISA, whereas Euroimmun IgA assay and Mikrogen IgA assay detected positive IgA levels from day 11 after onset of symptoms. Seroconversions for IgG occurred later than that for IgA and positive IgG levels were detected from day 11 on with the SERION IgG ELISA and the Mikrogen IgG assay, whereas the Euroimmun IgG assay failed to detect IgG antibody levels until day 24. Interestingly both IgA and IgG levels decreased after day 17 in all but the Euroimmun IgG assay, where the levels of detected antibodies even continued to increase. Neutralizing antibodies could be detected from day 11 on with a peak on day 14. The flow cytometry bead-based assay gave ambiguous results.

## Discussion

As a large variety of serological assays for the detection of anti-SARS-CoV-2 antibodies are now becoming available, proper assay validation is required to inform decisions on the use of these assays and the interpretation of test results for specific clinical and public health care demands. In this study we determined the specificity and sensitivity of three commercially available immunoassays and three rapid lateral flow tests for the detection of SARS-Cov-2 antibodies in comparison with a neutralization test. Neutralization assays remain the gold standard for determining antibody efficacy. 90 % of the PCR-confirmed COVID-19 patients investigated in this study showed neutralizing antibodies against the SARS-CoV-2 spike protein up to seven months after symptom onset. This is in line with literature data ranging from 77 % (19) to 96 % (30). Neutralizing antibody production seems to be correlated with severity of disease (30) and one study found neutralizing antibodies only in 12.5 % of asymptomatic SARS-CoV-2 positive patients (31). Both asymptomatic patients included in this study did not show neutralizing antibodies either, and it is open to be discussed. whether the original PCR results were false positive. The persistance of IgG antibodies has yet to be defined. However IgG levels seem to persist for at least seven months, as detected with different commercial assays as well as the bead-based surrogate test and the pseudovirus neutralisation assay. In comparison, SARS-CoV-1 patients were shown to maintain IgG antibodies for an average of two years, and significant reduction of IgG levels and titres occurred in the third year (32).

Sensitivity of the seven assays to detect sera with neutralizing activtity ranged from 7.0 % to 98.3 %. Specificity varied between 86.0 % and 100.0 %. Sensitivity was higher in IgG assays compared to IgM and IgA, and IgA levels were only negatively correlated with time from symptom onset in one assay. Based on the currently available data, IgM and IgG seroconversion occurs within 10 to 12 days and 12 to 14 days, respectively, after onset of symptoms (20, 33-37). A combination of IgA and IgM assays in SARS-CoV-2 diagnostics may increase sensitivity compared to a single IgG assay at the cost of a lower specificity depending on the assay. ELISAs were more sensitive compared to the lateral flow assays with lower sensitivity values for anti-SARS-CoV-2 IgG in our cohort compared to previously published data (14). Only the SERION ELISA agile SARS-CoV-2 IgG showed a satisfying sensitivity and specificity of more than 98 %. This is in line with data from the only published study on this assay (19). The bead-based surrogate test developed for unequivocally predicting neutralizing activity showed very high specificity and was the only surrogate test which reliably identified COVID-19 samples lacking neutralizing activity against the spike protein.

The symptoms most commonly described in this study were olfactory or gustatory dysfunction which occurred in 69 % of the participants. These data are in line with previous studies on this topic (38, 39).

### Limitations of the study are as follows

The severity of disease was not queried and one participant suffered from an immunodeficiency. Recruiting participants in a health care setting resulted in a gender imbalance towards the female gender. Though there are inter-assay differences in sensitivity, the number of participants was not high enough to assess a significant superiority. It is noteworthy that the three immune assays incorporated in this study are based on different antigen components. Whereas the Euroimmun assay is based on S1 antigen, Mikrogen *recomWell* is based on the nucleocapsid protein as antigen and SERION ELISA *agile* is coated with both S1 and S2 and the nucleocapsid. In the patient, followed over time IgG dynamics differ between the different assays.

Data from participants excluded from the main analysis whose COVID-19 disease was not confirmed by PCR, but by an unknown antibody assay, and who did not show neutralizing antibodies highlights the danger of COVID-19 diagnoses by antibody detection with apparently insufficient assays.

In this study only one assay (SERION ELISA agile SARS-CoV-2 IgG) showed a sensitivity and specificity of greater than 98 % and may be suitable for widespread assessment whether sera have neutralizing activity against SARS-CoV-2, for the clarification of doubtful PCR results and to determine if patients with potential post-COVID-19-symptoms have had an undetected COVID-19 disease as well as for epidemiological investigations.

## Data Availability

The data shown in the manuscript is available upon request from the corresponding author.

## Contribution

MK, TTL, CS, UV, AS, OK and ASU carried out the sample collection.

JG carried out the immunoassays and collected the questionnaire data.

SK, SB and GG established and carried out the (micro) neutralization tests.

NB and TK established the bead-based test.

FW and JWi produced recombinant RBD protein.

JWa, LD, OK and ASU contributed to the conception and design of the study.

MK and ASU analyzed the data.

OK raised study funding.

OK and ASU supervised the study.

MK and ASU wrote the first draft of the manuscript.

JG, JWa, TTL, CS, UV, AS, FW, JWi, TK, NB, SK, SB, LD, GG and OK reviewed and modified the manuscript and approved its final version.

## Acknowledgements

We thank Evgeny Golubtsov for sample collection support. The authors like to thank Florian Krammer, Icahn School of Medicine, Mount Sinai Hospital, New York, for providing the RBD expression plasmid, Gert Zimmer, Markus Hoffmann and Stefan Pöhlmann for providing reagents and protocols for pseudovirus generation, and Franziska Seifert for expert technical assistance with the bead-based assay.

